# The relationship between low levels of albuminuria and cardiovascular mortality among apparently healthy adults

**DOI:** 10.1101/2023.12.21.23300378

**Authors:** Sophie E Claudel, Sushrut S Waikar, Insa M. Schmidt, Ramachandran S. Vasan, Ashish Verma

**Author notes:** <u>Corresponding Author:</u> Ashish Verma, M.B.,B.S., 14 Evans Biomedical Research Center, X-521, 650 Albany Street, Boston University Chobanian & Avedisian School of Medicine, Boston, MA, 02118.

## Abstract

**Background:** Albuminuria is associated with cardiovascular events among adults with underlying cardiovascular disease and diabetes, even at low levels of urinary albumin excretion. We hypothesized that low levels of albuminuria in the ‘normal’ range (urinary albumin-to-creatine ratio (UACR) <30 mg/g) are associated with cardiovascular death among apparently healthy adults.

**Methods:** We studied adults who participated in the 1999-2014 National Health and Nutrition Examination Survey. We excluded participants with baseline cardiovascular disease, hypertension, diabetes, estimated glomerular filtration rate (eGFR) <60ml/min/1.73m^2^, those who were currently pregnant, and those who had received dialysis in the last year. After excluding these conditions, only 5.0% of the remaining population had UACR ≥30 mg/g (N=873) and were excluded. The final sample size was 16,247. We assessed the relationship between UACR and cardiovascular and all-cause mortality using multivariable-adjusted Cox proportional hazards models. Models were adjusted for age, sex, race or ethnicity, smoking status, systolic blood pressure, hemoglobin A1c, total cholesterol, health insurance, food insecurity, serum albumin, body mass index, use of statins, and eGFR.

**Results:** Mean age was 38.9 years (SD 13.6) and 53.7% were women. The median length of follow-up was 12.2 years. In multivariable-adjusted models, each doubling of UACR (within the <30 mg/g range) was associated with a 36% higher risk of cardiovascular death [HR 1.36 (95% confidence interval (CI) 1.11-1.65)] and a 28% higher risk of all-cause mortality [HR 1.28 (95%CI 1.17-1.41)]. The highest tertile of UACR (7.1-29.9 mg/g) was associated with an 87% higher risk of cardiovascular death [HR 1.87 (95%CI 1.20-2.92)] and 59% higher risk of all-cause mortality [HR 1.59 (95%CI 1.28-1.96)], compared with the lowest tertile (< 4.3 mg/g).

**Conclusions:** In a nationally representative sample of relatively healthy community-dwelling adults, higher levels of albuminuria in the conventionally “normal” range <30 mg/g in healthy individuals are associated with greater mortality. Overall, our findings contribute to the growing body of evidence on the existence of a risk gradient across all levels of albuminuria, even in the so-called normal range.

## Introduction

Healthy kidneys filter over 100 liters of plasma water daily to excrete waste products into a few liters of urine, which are virtually free of protein.^1^ Persistent proteinuria indicates a pathological state. Albuminuria, specifically, is often attributed to glomerular pathology (such as podocyte effacement in the setting of diabetes mellitus) but can also be due to failure of proximal tubular catabolism and reabsorption.^1,2^ A key element in the diagnosis of chronic kidney disease (CKD) is elevated urinary albumin excretion, which is defined as a urinary albumin-to-creatinine ratio (UACR) above 30 mg/g.^3^ CKD is a well-established risk factor for cardiovascular disease (CVD) and cardiovascular mortality.^4^ However, increasing evidence points to cardiovascular risk associated with albuminuria below this threshold.^5–7^

Prior work has demonstrated an association between albuminuria and incident CVD (such as myocardial infarction, coronary revascularization, and heart failure hospitalization),^5^ incident heart failure,^7^ and left-ventricular hypertrophy at values below the threshold of UACR < 30 mg/g.^6^These studies have been predominantly performed among individuals with known CVD risk factors, including hypertension and diabetes. It is unknown whether albuminuria in a “low-grade” range (UACR < 30 mg/g) is clinically meaningful in healthy adults without pre-existing cardiovascular risk factors.

We hypothesized that higher levels of albuminuria within the conventional clinical normal range (UACR < 30 mg/g) would be associated with graded risk of cardiovascular and all-cause mortality among relatively healthy adults without major CVD risk factors.

## Methods

We included 16,247 adults participating in the National Health and Nutrition Examination Survey (NHANES) between 1999 and 2014.^8^ We excluded participants who were <20 years old, had hypertension (defined by systolic blood pressure >130 mmHg, diastolic blood pressure >80 mmHg, or use of antihypertensive medications), had diabetes (defined by hemoglobin A1c (HbA1c) ≥6.5% or use of hypoglycemic agents), had prevalent CVD (self-reported myocardial infarction, heart disease, heart failure, angina, or stroke), were pregnant, or had an estimated glomerular filtration rate (eGFR) <60 ml/min per 1.73m^2^ (**Figure 1**). After excluding these conditions, only 5.0% of the remaining population had UACR ≥30 mg/g (N=873) and 1% (N=180) were missing UACR, which was excluded. The final sample size was 16,247. The 1999-2000 and 2005-2006 serum creatinine values were re-calibrated with a modified Deming equation per NHANES documentation, for consistency across survey years. eGFR was calculated using the 2021 CKD-EPI creatinine-based equation.^9^

**Figure 1.**
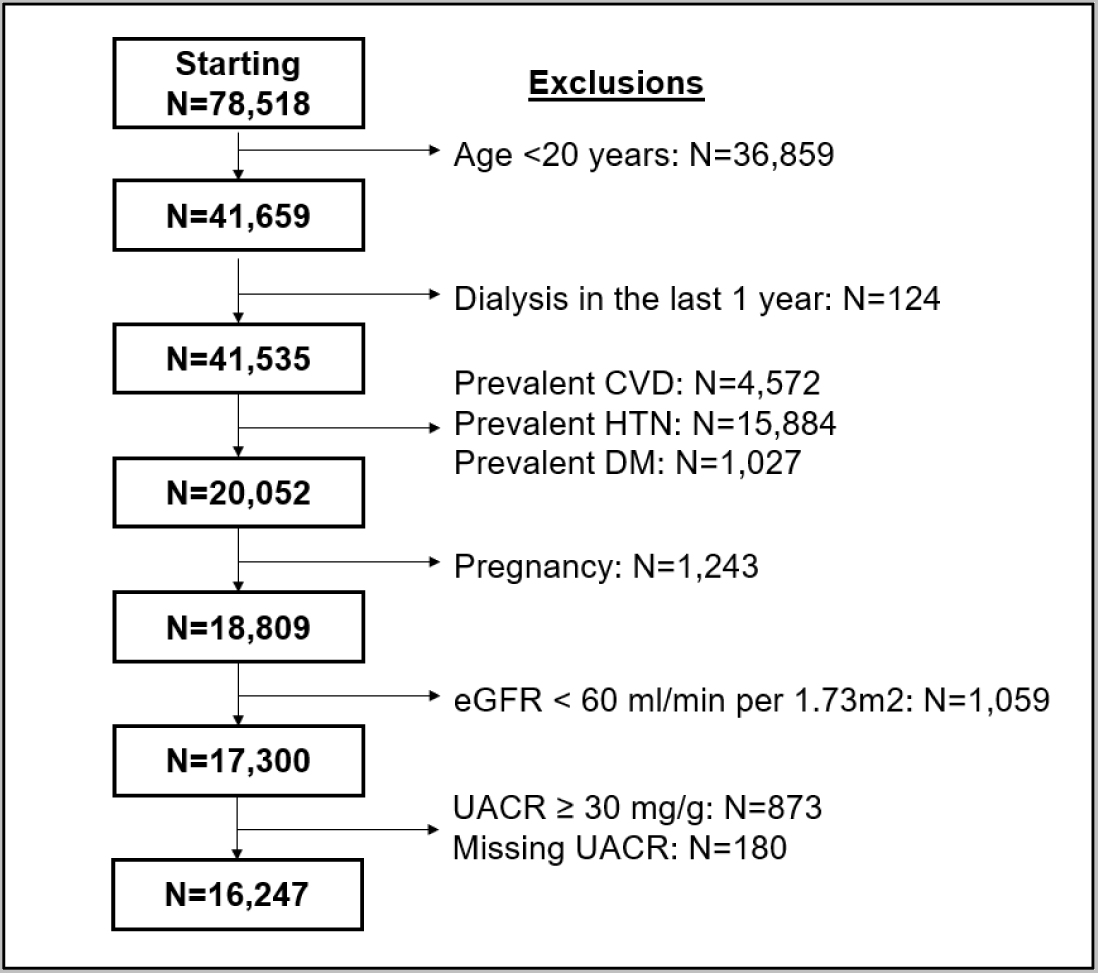
Sample selection. Abbreviations: CVD, cardiovascular disease; DM, diabetes mellitus; eGFR, estimated glomerular filtration rate; HTN, hypertension; UACR, urinary albumin-to-creatinine ratio.

### Exposure variable

UACR was calculated based on measured urinary albumin (solid-phase fluorescent immunoassay) and urinary creatinine (Jaffe reaction). The lowest value for UACR was 0.12 mg/g.

### Outcome variables

We linked the participants to the National Death Index and assessed cardiovascular and all-cause mortality. Cardiovascular mortality included death from heart disease (ICD-10 codes I00-I09, I11, I13, I20-I151) and cerebrovascular diseases (ICD-10 codes I60-I69). A sensitivity analysis assessed the association with non-cardiovascular mortality.

### Covariates

We included the following covariates based on biological and clinical plausibility: age (years), sex (male, female), race or ethnicity (non-Hispanic White, Mexican American, other Hispanic, non-Hispanic Black, other), smoking status (never, former/current), systolic blood pressure (mmHg), HbA1c (%), total cholesterol (mg/dL), health insurance, food insecurity, serum albumin (mg/dL), body mass index (BMI, kg/m^2^), use of statin, and eGFR (ml/min per 1.73m^2^).

### Statistical analysis

We first summarized the sample characteristics using weighted sample mean (standard deviation (SD)), median [interquartile range (IQR)], or weighted percentage. Chi-squared testing compared weighted proportions across tertile of UACR and univariate linear regression with contrast estimates compared means. Total missing data was less than 7% (**Table S1**), therefore, all subsequent analyses were done using complete case analysis.

We used linear regression modeling to assess the linear associations between participant characteristics and albuminuria, using natural logarithmically transformed UACR. We then sequentially added hypothesized predictors to a multivariable model and retained only predictors with *p*<0.10. We performed a sensitivity analysis repeating this with the dependent variable as urine albumin alone, not the ratio with urinary creatinine, and adjusting for urine creatinine. Beta coefficients were then expressed as percent change.

We used Cox proportional hazard regression models to examine the multivariable-adjusted risk of outcomes according to UACR modeled as a continuous variable (logarithm base 2) and tertiles. We tested the assumption of proportionality of hazards with Schoenfeld residuals. The linearity assumption was checked using restricted cubic splines using the ‘rms’ package in R software. The multivariable adjustment strategy for the Cox proportional hazard models was based on covariates’ biological and clinical plausibility as potential confounders of the association between UACR and CVD mortality and all-cause mortality. Model 1 was adjusted for age, sex, self-reported race or ethnicity, insurance, smoking status, BMI, and survey year. Model 2 was additionally adjusted for HbA1c, total cholesterol, systolic blood pressure, serum albumin, and statin use. Model 3 was additionally adjusted for eGFR.

We performed sensitivity analyses using the exposure variable of urinary albumin and adjusting for urinary creatinine as a covariate, as well as assessing the association between UACR and non-cardiovascular mortality. We performed additional sensitivity analyses using dichotomized UACR (≥7 mg/g) as the exposure variable as well as looking at UACR scaled per 10 mg/g.

Kaplan-Meier curves were used to visualize the survival probability by tertile of UACR and a log-rank test compared estimates of the hazard function for each tertile.

All analyses were formed in SAS (version 9.4, Cary, NC); Kaplan-Meier curves were generated in GraphPad Prism (version 10.0, Boston, MA) and splines were plotted in R software (version 3.5). Statistical significance was determined by *p*<0.05. All data are publicly available for download at https://wwwn.cdc.gov/nchs/nhanes/default.aspx.

## Results

The mean age of the sample was 38.9 years (SD 13.6), 53.7% of the sample were women, and 68.5% were non-Hispanic White (**Table 1**). The median UACR was 5.3 mg/g [IQR 3.7, 8.1] overall; The UACR range in each tertile was 0.1-4.3 mg/g, 4.3-7.1 mg/g, and >7.1mg/g. Participant characteristics by tertile of UACR are shown in **Table 1**. In general, participants in the highest UACR tertile were slightly older, more frequently women, and were more likely to experience food insecurity than those in tertile 1. Other clinical and demographic characteristics were similar across tertiles of UACR. HbA1c ranged from 3.3 to 6.4% (mean 5.3%, SD 0.3).

**Table 1.** Participant characteristics.

| **Table 1. Participant characteristics** |  |  |  |  |  |
| --- | --- | --- | --- | --- | --- |
|  | **Overall** | **Tertile 1**<br>**UACR < 4.3 mg/g** | **Tertile 2**<br>**UACR 4.3-7.1 mg/g** | **Tertile 3**<br>**UACR > 7.1 mg/g** | **p-value** |
| No. of participants | 16,247 | 5,422 | 5,409 | 5,416 |  |
| UACR, median [IQR] | 5.3 [3.7, 8.1] | 3.3 [2.7, 2.8] | 5.4 [4.8, 6.1] | 10.3 [8.4, 14.2] |  |
| Age, years | 38.9 (13.6) | 37.8 (12.3) | 39.5 (13.2) | 40.5 (15.0) | <0.0001 |
| Female sex, % | 53.7 | 36.6 | 57.7 | 68.6 | <0.0001 |
| Race or Ethnicity, % |  |  |  |  | <0.0001 |
| Non-Hispanic White | 68.5 | 69.0 | 69.0 | 67.5 |  |
| Mexican American | 9.7 | 8.3 | 9.8 | 10.9 |  |
| Other Hispanic | 6.4 | 5.9 | 6.8 | 6.6 |  |
| Non-Hispanic Black | 9.2 | 11.3 | 7.9 | 8.4 |  |
| Other Race | 6.2 | 5.5 | 6.5 | 6.6 |  |
| Education, % |  |  |  |  | 0.006 |
| Below High School | 15.8 | 14.5 | 15.7 | 17.5 |  |
| High School | 22.1 | 22.4 | 22.5 | 21.3 |  |
| Above High School | 62.1 | 63.1 | 61.8 | 61.2 |  |
| Income |  |  |  |  | <0.0001 |
| Low income | 14.5 | 13.2 | 13.9 | 16.5 |  |
| Middle income | 34.1 | 32.0 | 34.5 | 36.0 |  |
| High income | 51.4 | 54.8 | 51.6 | 47.5 |  |
| Uninsured, % | 23.1 | 22.3 | 23.5 | 23.4 | 0.40 |
| Food insecure, % | 13.0 | 12.0 | 12.8 | 14.3 | 0.005 |
| Smoking status, % |  |  |  |  | 0.23 |
| Never | 55.4 | 54.7 | 55.3 | 56.2 |  |
| Former | 19.3 | 19.3 | 20.2 | 18.4 |  |
| Current | 25.3 | 26.0 | 24.6 | 25.3 |  |
| Systolic blood pressure, mmHg | 112.2 (8.9) | 112.6 (8.6) | 112.0 (9.0) | 112.2 (9.2) | 0.01 |
| Diastolic blood pressure, mmHg | 67.4 (7.8) | 67.4 (7.9) | 67.4 (7.7) | 67.3 (7.8) | 0.79 |
| Hemoglobin A1c, % | 5.3 (0.3) | 5.2 (0.3) | 5.3 (0.3) | 5.3 (0.4) | <0.0001 |
| BMI, kg/m<sup>2</sup> | 27.1 (5.7) | 27.1 (5.2) | 27.0 (5.7) | 26.7 (6.2) | <0.0001 |
| LDL, mg/dl | 116.3 (23.3) | 116.7 (23.3) | 117.1 (23.8) | 114.7 (22.8) | 0.08 |
| Total cholesterol, mg/dl | 193.6 (37.8) | 193.2 (38.2) | 194.9 (39.6) | 192.9 (38.4) | 0.08 |
| eGFR, ml/min/1.73m<sup>2</sup> | 102.8 (16.5) | 100.4 (16.2) | 104.0 (16.0) | 104.4 (17.0) | <0.0001 |
| Statin use, % | 4.3 | 3.3 | 4.5 | 5.3 | 0.0003 |
| p-values were determined by Chi-squared testing (for weighted proportions) or univariate linear regression with contrast estimates (for means). |  |  |  |  |  |
| Abbreviations: BMI, body mass index; eGFR, estimated glomerular filtration rate; GED, general education diploma; LDL, low density lipoprotein; UACR, urine albumin-to-creatinine ratio. |  |  |  |  |  |

### Clinical correlates of albuminuria below 30 mg/g

Multivariable adjusted associations between participant characteristics and UACR are shown in **Table 2**. The strongest clinical correlate of UACR was HbA1c. Every 1% increase in HbA1c was associated with a 9.4% increase in UACR (9.4%, 95% CI 5.7 to 13.1, *p*<0.0001). Additional clinical correlates associated with increased UACR included food insecurity (% change: 5.0%, 95% CI 1.7 to 8.3, *p*=0.003), lack of health insurance (% change: 3.8%, 95% CI 0.8 to 6.8, *p*=0.01), and systolic blood pressure (% change: 2.9% for every 10 mmHg increase, 95% CI 1.3 to 4.6, *p*=0.0004). Several characteristics were associated with lower UACR, including male sex (% change: −30.6%, 95% CI −32.2 to −28.8, *p*<0.0001), eGFR (% change: −7.1% for every 10 ml/min/1.73m^2^ decrease, 95% CI −8.1 to −6.2, *p*<0.0001), self-reported non-Hispanic Black race (% change: −4.4%, 95% CI −7.6 to −1.1, *p*=0.009), and higher BMI (% change: −2.1% for every 5 kg/m^2^, 95% CI −3.1 to −1.1, *p*<0.0001). We performed an additional analysis using urine albumin concentration (not the ratio) as the dependent variable and controlling for urine creatinine concentration, which resulted in similar findings (**Table S2**).

**Table 2.** Clinical correlates of low-grade albuminuria.

|  | Univariate |  |  | Multivariable |  |  |
| --- | --- | --- | --- | --- | --- | --- |
| | $\beta$ Coefficient<br>(95% CI) | % Change<br>(95% CI) | *p*-value | $\beta$ Coefficient<br>(95% CI) | % Change<br>(95% CI) | *p*-value |
| 10-year increase in age | 0.04 (0.04, 0.05) | 4.0 (3.1, 4.9) | <0.0001 | 0.09 (0.07, 0.1) | 9.3 (8.1, 10.6) | <0.0001 |
| Male sex | -0.3 (-0.3, -0.3) | -27.0 (-28.6, -25.4) | <0.0001 | -0.4 (-0.4, -0.3) | -30.6 (-32.2, -28.8) | <0.0001 |
| Race or Ethnicity |  |  |  |  |  |  |
| Non-Hispanic White | [Ref] | [Ref] |  | [Ref] | [Ref] |  |
| Mexican | 0.05 (0.01, 0.08) | 4.9 (1.2, 8.6) | 0.008 | -0.009 (-0.05, 0.03) | -0.9 (-4.6, 2.8) | 0.62 |
| Other Hispanic | 0.001 (-0.04, 0.04) | 0.1 (-4.1, 4.6) | 0.95 | -0.04 (-0.08, 0.004) | -3.8 (-7.8, 0.4) | 0.08 |
| Non-Hispanic Black | -0.1 (-0.1, -0.06) | -9.1 (-12.2, -5.8) | <0.001 | -0.05 (-0.08, -0.01) | -4.4 (-7.6, -1.1) | 0.009 |
| Other | 0.03 (-0.02, 0.07) | 3.0 (-1.5, 7.8) | 0.20 | -0.002 (-0.05, 0.04) | -0.2 (-4.7, 4.4) | 0.92 |
| Education |  |  |  |  |  |  |
| Below High School | 0.05 (0.01, 0.08) | 4.7 (1.5, 8.1) | 0.004 | 0.01 (-0.03, 0.03) | 1.5 (-2.0, 5.1) | 0.40 |
| High School / GED | 0.002 (-0.03, 0.03) | 0.2 (-2.7, 3.1) | 0.91 | -0.002 (-0.03, 0.03) | -0.2 (-3.0, 2.7) | 0.88 |
| Above High School | [Ref] | [Ref] |  | [Ref] | [Ref] |  |
| Uninsured | 0.007 (-0.02, 0.03) | 0.7 (-2.0, 3.5) | 0.60 | 0.04 (0.008, 0.07) | 3.8 (0.8, 6.8) | 0.01 |
| Food insecurity | 0.05 (0.02, 0.08) | 4.8 (1.8, 7.9) | 0.002 | 0.05 (0.02, 0.08) | 5.0 (1.7, 8.3) | 0.003 |
| Current/former smoking | -0.01 (-0.03, 0.01) | -1.1 (-3.2, 1.1) | 0.32 | - | -- |  |
| 5kg/m<sup>2</sup> increase in BMI | -0.02 (-0.03, -0.01) | -2.3 (-3.3, -1.3) | <0.0001 | -0.02 (-0.03, -0.01) | -2.1 (-3.1, -1.1) | <0.0001 |
| 1% increase in hemoglobin A1c | 0.09 (0.06, 0.1) | 9.0 (5.8, 12.2) | <0.0001 | 0.09 (0.06, 0.1) | 9.4 (5.7, 13.1) | <0.0001 |
| 1mg/dL increase in serum albumin | -0.004 (-0.008, -0.0004) | -0.4 (-0.8, -0.04) | 0.03 | 0.02 (0.01, 0.02) | 1.9 (1.5, 2.3) | <0.0001 |
| 50mg/dL increase in total cholesterol | -0.0008 (-0.02, 0.01) | -0.1 (-1.5, 1.4) | 0.91 | -0.03 (-0.04, -0.01) | -2.8 (-4.2, -1.4) | 0.0002 |
| 10ml/min decrease in eGFR | -0.03 (-0.04, -0.03) | -3.4 (-4.1, -2.7) | <0.001 | -0.07 (-0.08, -0.06) | -7.1 (-8.1, -6.2) | <0.0001 |
| 10mmHg increase in SBP | -0.02 (-0.03, -0.002) | -1.8 (-3.3, -0.2) | 0.02 | 0.03 (0.01, 0.04) | 2.9 (1.3, 4.6) | 0.0004 |
| 5mmHg increase in DBP | -0.007 (-0.01, 0.001) | -0.7 (-1.4, 0.1) | 0.11 | - | -- |  |

Smoking status and DBP not included in multivariable regression as $p > 0.10$ when added to the model.
Percent change reflects exponentiated $\beta$ coefficients.
Abbreviations: BMI, body mass index; CI, confidence interval; DBP, diastolic blood pressure; eGFR, estimated glomerular filtration rate; GED, general education diploma; SBP, systolic blood pressure.

### Association between UACR and mortality

Over a median follow-up of 12.2 years, 177 individuals died from CVD, 777 from non-cardiovascular causes, and 954 from all causes. The results for the association between UACR and risk of mortality are shown in **Table 3**. In multivariable-adjusted models, each doubling of UACR was associated with a 36% higher risk of cardiovascular death (HR 1.36, 95% CI 1.11-1.65, *p*=0.003) and a 28% higher risk of all-cause mortality (HR 1.28, 95% CI 1.17-1.41, *p*<0.0001). Individuals with UACR between 7.1 and 29.9 mg/g had an 87% higher risk of cardiovascular death (HR 1.87, 95% CI 1.20-2.92) and 59% higher risk of all-cause mortality (HR 1.59, 95% CI 1.28-1.96) than those with UACR below 4.3 mg/g. Similar but attenuated associations were found for non-cardiovascular mortality (**Table S3**).

**Table 3.**
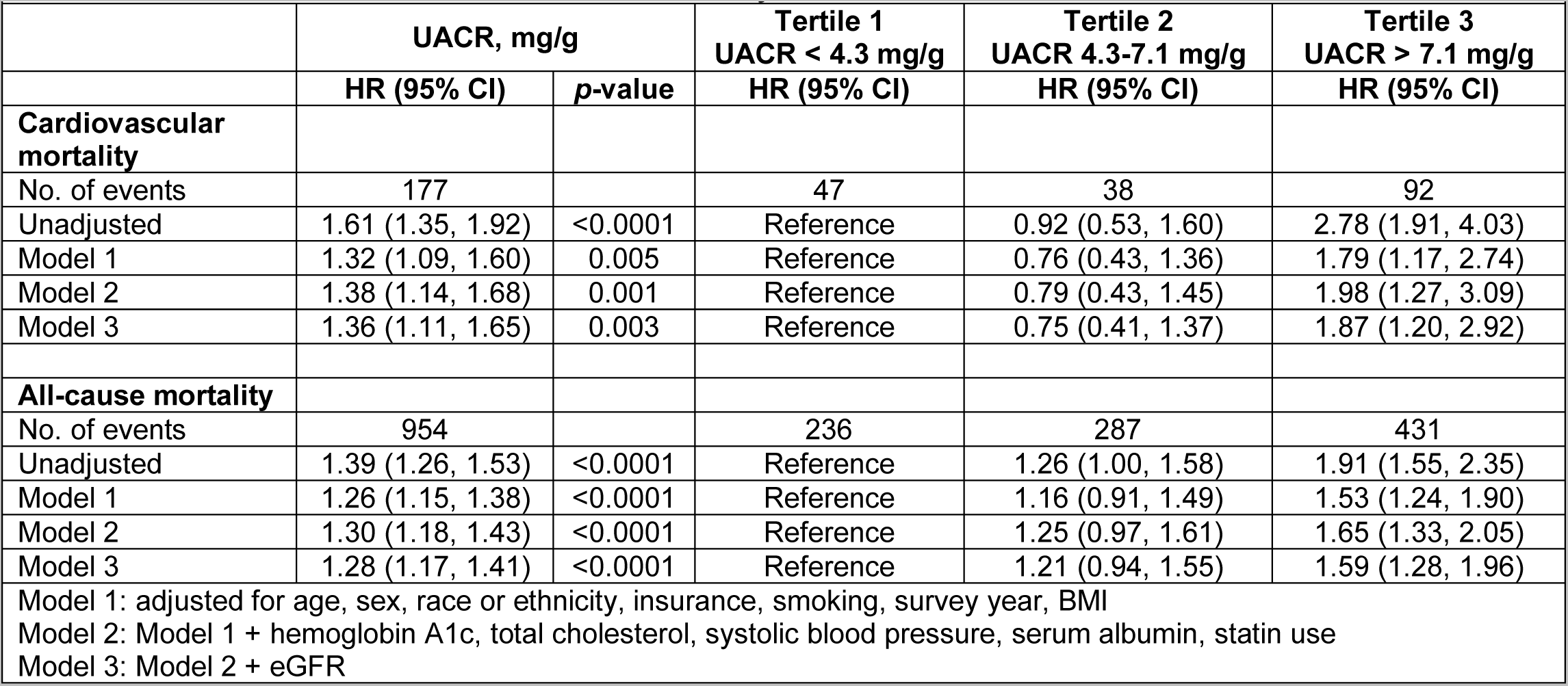
Association between albuminuria and mortality.

Kaplan-Meier curves demonstrate decreased survival among participants in the highest tertile of UACR (7.1 to 29.9 mg/g) for cardiovascular mortality and all-cause mortality (log-rank test *p*<0.0001; **Figure 2**).

**Figure 2.**
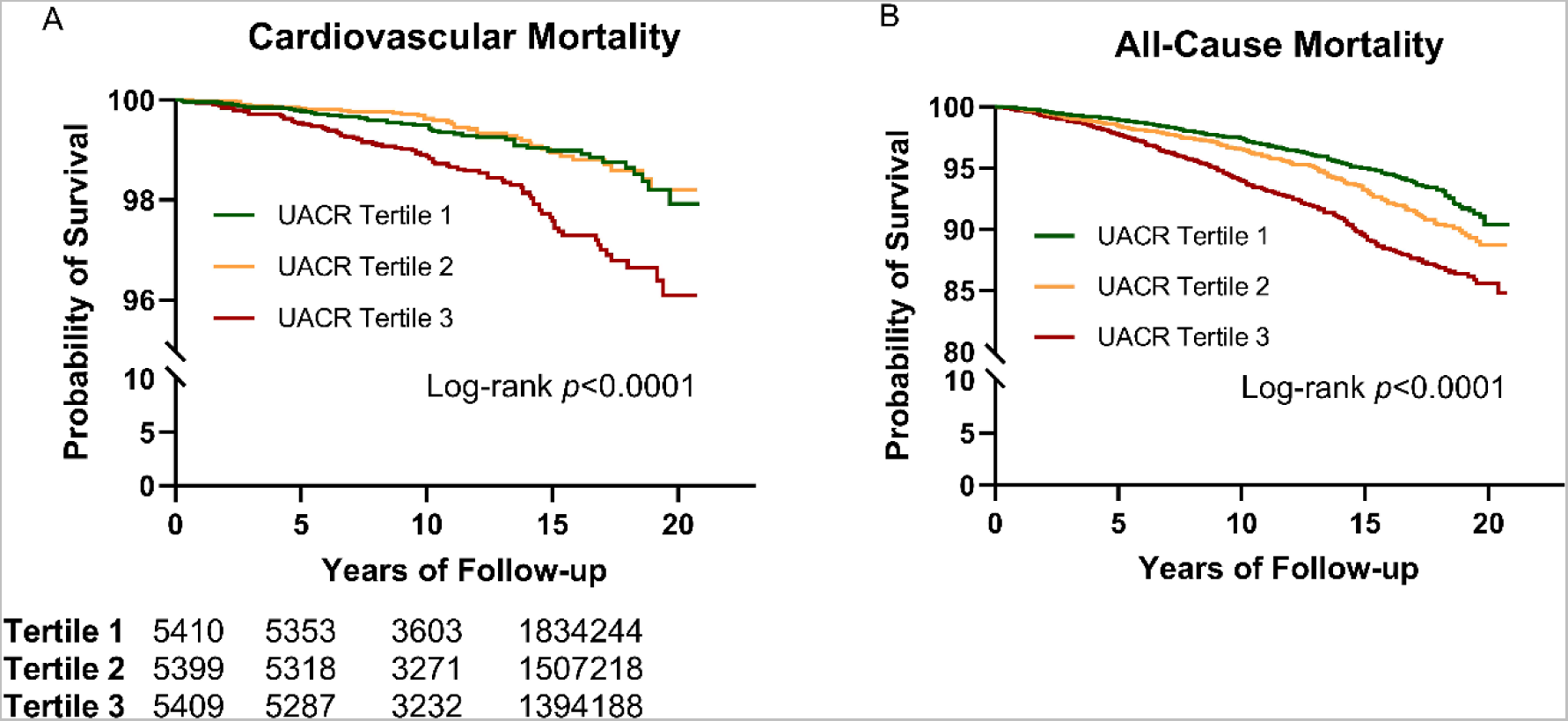
Survival probability by tertile of UACR. Kaplan-Meier curves of a) cardiovascular mortality and b) all-cause mortality, by tertile of UACR. UACR values range from 0.1-4.3 mg/g for tertile 1, 4.3-7.1 mg/g for tertile 2, and 7.1-29.9 mg/g for tertile 3. Y-axis breaks included for improved visualization of the curves.

Figure 3 shows the continuous relationship between UACR and risk of CVD death and all-cause mortality in the multivariable-adjusted model (*p*-value for linearity <0.0001). The spline analysis demonstrated that the HR for each mortality outcome exceeded 1.0 at a UACR of 7 mg/g (Figure 3).

**Figure 3.**
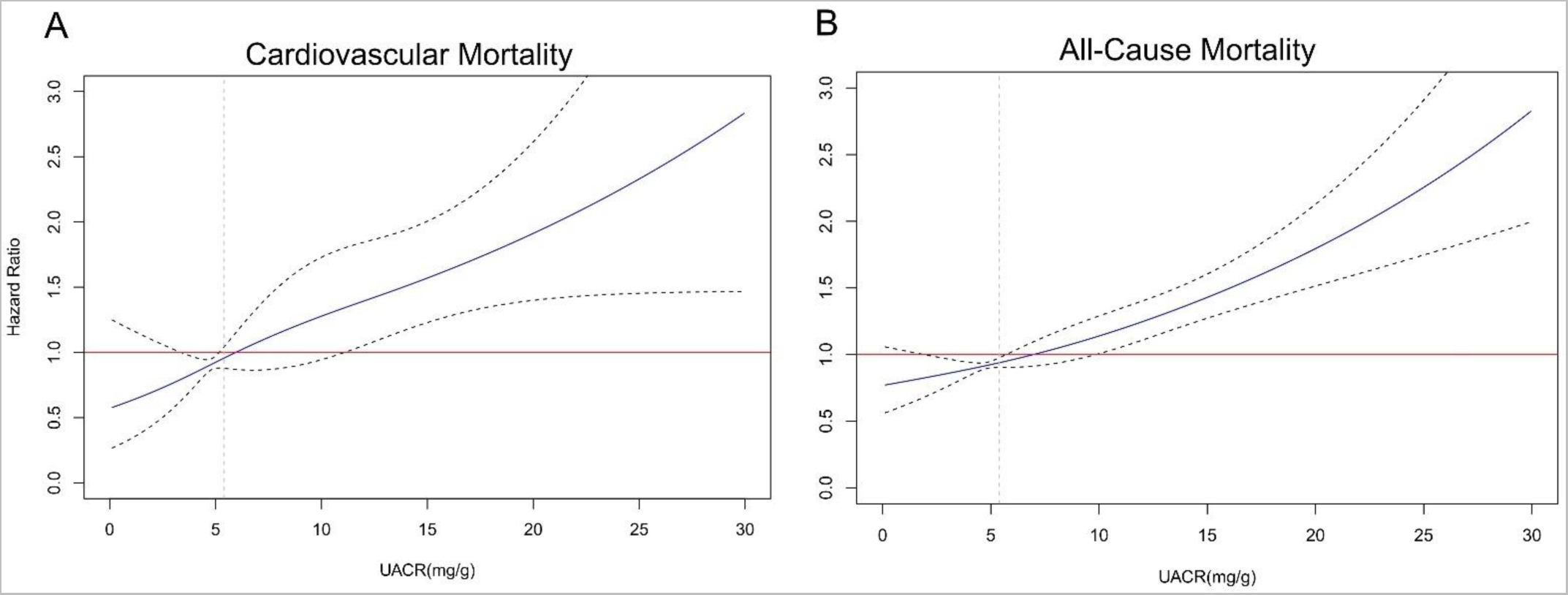
Restricted cubic splines. Linear relationship between UACR and (a) cardiovascular mortality and (b) non-cardiovascular mortality. The *p*-value for linearity is <0.0001 for all. Models are adjusted for age, sex, race or ethnicity, insurance, smoking, survey year, BMI, hemoglobin A1c, total cholesterol, systolic blood pressure, serum albumin, statin use, and eGFR.

The findings remained qualitatively similar in the sensitivity analysis when using urinary albumin as the outcome variable and adjusting for urinary creatinine (Figure 4), when modeling the exposure (UACR) as a dichotomized variable at the level of 7 mg/g, and when scaled per 10 mg/g (**Table S4**). There was no evidence of effect modification by sex, race or ethnicity, HbA1c, or BMI for cardiovascular mortality; however, there was a significant interaction with BMI for both non-cardiovascular and all-cause mortality (Figure 5, **Table S5**).

**Figure 4.**
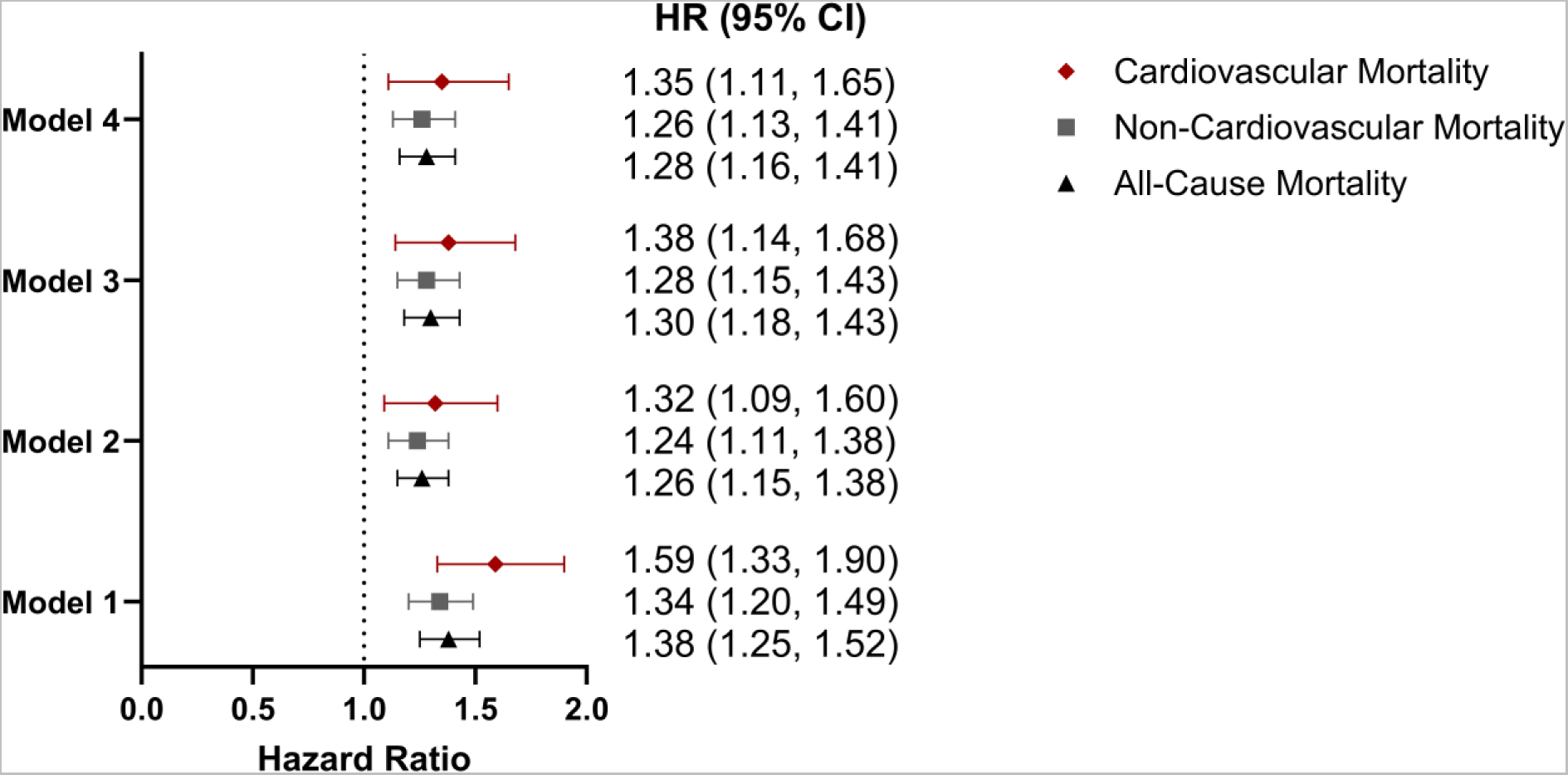
The association between urinary albumin alone (not the ratio) and mortality, adjusting for urinary creatinine. Hazard ratio represents a doubling of urinary albumin concentration. Model 1 is adjusted for urinary creatinine (log_2_-transformed). Model 2 is further adjusted for age, sex, race or ethnicity, insurance, smoking, survey year, and BMI. Model 3 is further adjusted for hemoglobin A1c, total cholesterol, systolic blood pressure, serum albumin, and statin use. Model 4 is additionally adjusted for eGFR.

**Figure 5.**
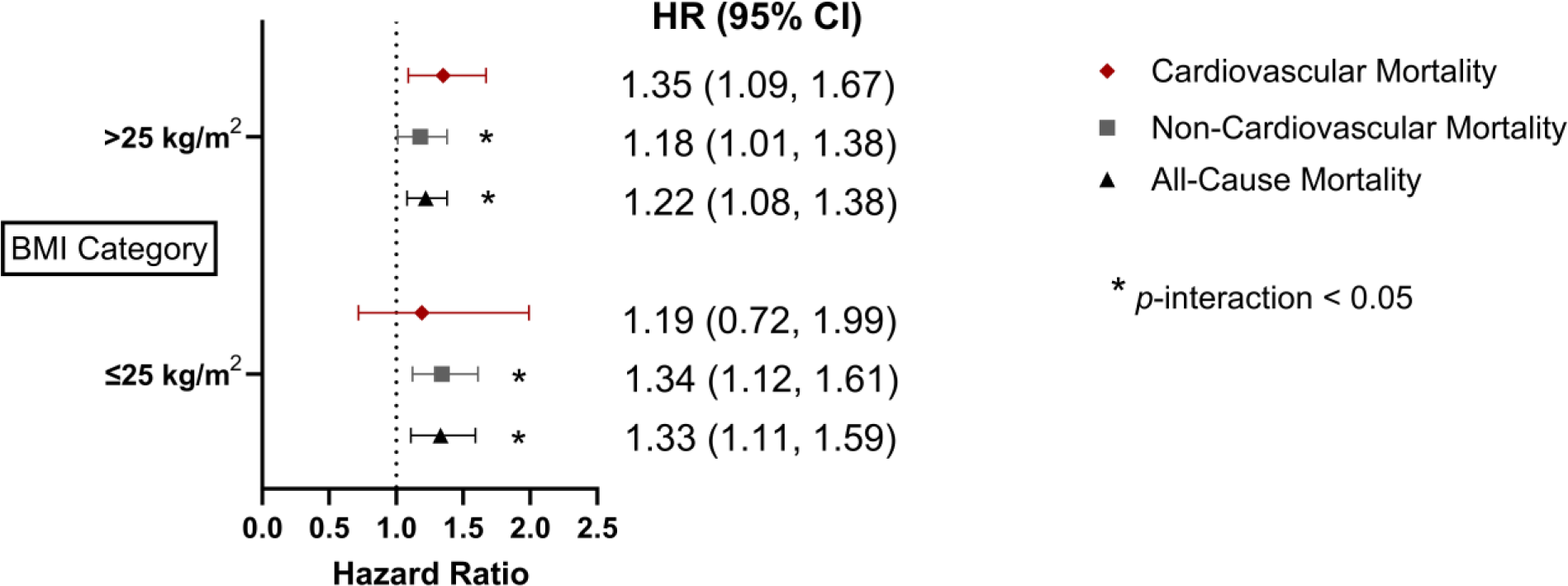
Association between UACR and mortality, stratified by BMI. Hazard ratios represent a doubling in UACR. Models adjusted for age, sex, race or ethnicity, insurance, smoking, survey year, BMI, hemoglobin A1c, total cholesterol, systolic blood pressure, serum albumin, statin use, and eGFR.

## Discussion

In our investigation, albuminuria within the range from 0.12 to 30 mg/g (conventionally considered “normoalbuminuria”) displayed a graded relationship with cardiovascular mortality risk within a nationally representative sample of U.S. adults without major CVD risk factors. These findings challenge the notion that all UACR values below 30 mg/g represent non-pathological albumin excretion and provide further support for albuminuria as a potential, unique CVD risk factor across levels conventionally considered normal. The novelty of these analyses is in the use of a relatively healthy population, whereas prior studies of albuminuria and mortality were limited to older adults with prevalent CVD or known CVD risk factors.^10–14^

Our study extends the literature on the cardiovascular implications of albuminuria within the normal range by showing these findings in a large and diverse sample and with a long follow-up period.^15–18^ By examining UACR as a continuous exposure, rather than as a categorical variable,^15–18^ we were able to assess the degree of albuminuria at which mortality risk begins to rise. Current guidelines define albuminuria below 30 mg/g as ‘normal to mildly increased’ and focus treatment recommendations on more severe levels of albuminuria.^19^ This definition of ‘normal to mildly increased’ is based on a study of 609 Danish adults with diabetes in which those with UACR ≥ 30 μg per milliliter had a higher risk of progressing to more advanced proteinuria (>0.4g in spot morning urine sample) and mortality during follow up.^20^ While this aforementioned study was not intended to establish normal albuminuria ranges, the results have been widely extrapolated for this purpose. However, prior literature demonstrates the prognostic value of UACR values below 30 mg/g, because evidence of cardiovascular dysfunction (based on pulse-wave velocity, left-ventricular mass) increases at values as low as UACR 6.45 to 9.37 mg/g.^21,22^ Our analyses further support these findings as the splines demonstrated an increase in mortality once UACR reached approximately 7 mg/g. To date, there are no randomized trials assessing clinical outcomes of treating albuminuria in this range among patients at otherwise low risk of CVD. Nonetheless, our findings demonstrate the prognostic importance of an elevated UACR within the ‘so-called normal’ range.

A prior analysis of the association between albuminuria within the normal range and mortality in NHANES did so in the general adult population with cardiovascular comorbidities.^11^ The analysis included participants with prevalent CVD, hypertension, diabetes, impaired eGFR, or were pregnant or had previously received dialysis.^11^ Here, we present a population restricted to those without known clinical CVD or comorbidities that might affect either UACR or cardiovascular risk. The estimates therefore reflect a more traditionally ‘healthy’ population of U.S. adults and provide more direct evidence of the associations of UACR below 30 mg/g. Other studies have examined thresholds or quartiles of UACR below 30 mg/g without assessing the continuous relationship.^17,18^

There are several possible explanations for the relationship between albuminuria within the low-grade range and mortality. Low levels of albuminuria are a known risk factor for CKD,^23^ which itself is a risk factor for cardiovascular death.^4,24^ Therefore, adults with low-normal albuminuria may be at risk of CVD mortality through a pathway mediated by albuminuria progression and target organ damage to the vascular endothelial tissue and kidneys (i.e., incident CKD). ^25^ Alternatively, elevated albuminuria within the normal range may reflect low levels of clinically undetectable vascular endothelial damage (e.g., the Steno hypothesis^26,27^) and intravascular congestion, which subsequently result in cardiac strain.^28,29^ Supporting this hypothesis, impaired retinal arteriolar dilatory capacity – a direct measure of endothelial dysfunction – was associated with increasing albuminuria in adults with and without diabetes.^30^ Clinically, low-normal albuminuria was associated with incident hypertension among Korean adults participating in a health screening program.^15^ Differences in tubular reabsorption capacity may underly the reason for greater versus lower albuminuria; if this is the case, then the reason for differences in tubular uptake (e.g., due to genetic polymorphisms, tubular injury, or other mechanism) may account for the observed association. Any of these, or a combination of mechanisms, may account for the association between albuminuria and cardiovascular mortality.^5,18,31,32^ The relationship between albuminuria and non-cardiovascular mortality may be secondary to systemic inflammation, which has previously been hypothesized to drive the relationship between albuminuria and incident cancer and cancer-associated mortality.^33,34^

Our analysis of clinical correlates of albuminuria revealed that food insecurity, lack of health insurance, higher HbA1c (but still below 5.4%), and higher systolic blood pressure (but still below 130 mmHg) were associated with increased UACR. Prior work has shown that the food environment may be a modifiable risk factor for CKD, as food insecurity is associated with a higher risk of end-stage kidney disease.^35,36^ Food insecurity may lead to the development of risk factors for albuminuria such as pre-diabetes, psychosocial distress, dietary acid load (lower consumption of fruits and vegetables), metabolic dysfunction associated steatotic liver disease, and elevated blood pressure.^35,37,38^ The seemingly paradoxical inverse associations between male sex, non-Hispanic Black race, and BMI with UACR likely due to differences in creatinine generation rate and urinary creatinine excretion. Prior work has attributed the seemingly paradoxical relationship between higher BMI and lower UACR to non-linear associations.^39^

Our investigation has several strengths, including a robust sample of healthy adults that reflect the overall U.S. population, long length of follow-up, and incorporation of non-traditional CVD risk factors such as food security into the models. However, we must note the limitations associated with the study design. Foremost, these analyses were based on a single measurement of UACR and, therefore, cannot account for within-person variability in albuminuria between repeated measures.^40,41^ Additionally, mortality outcomes were obtained from administrative data based on ICD-10 codes, which are prone to misclassification bias. Finally, a history of prior cardiovascular events was self-reported. Future analyses would benefit from the addition of vascular imaging modalities, such as pulse-wave velocity, carotid stiffness, and echocardiography, to assess the pathways between and mediators of UACR and CVD risk.^21,22,42^

In conclusion, in a nationally representative sample of apparently healthy community-dwelling adults, higher levels of albuminuria in the “normal” range <30 mg/g in healthy individuals are strongly associated with mortality. These findings demonstrate the prognostic value of UACR and highlight its potential as a CVD biomarker even among otherwise healthy adults. Our findings support the 2021 European Society of Cardiology recommendations for routine albuminuria screening in higher risk adults.^43^ Multiple new anti-proteinuric medications and weight loss therapies are now available; early implementation may prevent progressive, irreversible changes to the kidney and adverse CVD events within this population.^25^ Further work is needed to determine the optimal UACR threshold for initiating anti-proteinuric or other cardiometabolic/kidney therapies, as well as assess the cost-effectiveness of albuminuria screening in the general U.S. population.

## Data Availability

Data are publicly available at https://wwwn.cdc.gov/nchs/nhanes/default.aspx.

## References

1. Rose B. Pathophysiology of Renal Disease. 2nd edition. New York: McGraw Hill; 1987.

2. Dickson LE, Wagner MC, Sandoval RM, Molitoris BA. The proximal tubule and albuminuria: Really! Journal of the American Society of Nephrology 2014;25:443–453.

3. Kidney Disease: Improving Global Outcomes (KDIGO) Guideline Development Work Group. KDIGO 2012 Clinical Practice Guideline for the Evaluation and Management of Chronic Kidney Disease. Kidney Int Suppl (2011) 2013;3:136–150.

4. Jankowski J, Floege J, Fliser D, Böhm M, Marx N. Cardiovascular Disease in Chronic Kidney Disease Pathophysiological Insights and Therapeutic Options. Circulation 2021;143:1157–1172.

5. Ruggenenti P, Porrini E, Motterlini N, Perna A, Ilieva AP, Iliev IP, Dodesini AR, Trevisan R, Bossi A, Sampietro G, Capitoni E, Gaspari F, Rubis N, Ene-Iordache B, Remuzzi G. Measurable urinary albumin predicts cardiovascular risk among normoalbuminuric patients with type 2 diabetes. Journal of the American Society of Nephrology 2012;23:1717–1724.

6. Wang T, Zhong H, Lian G, Cai X, Gong J, Ye C, Xie L. Low-Grade Albuminuria Is Associated with Left Ventricular Hypertrophy and Diastolic Dysfunction in Patients with Hypertension. Kidney Blood Press Res 2019;44:590–603.

7. Blecker S, Matsushita K, Kttgen A, Loehr LR, Bertoni AG, Boulware LE, Coresh J. High-normal albuminuria and risk of heart failure in the community. American Journal of Kidney Diseases 2011;58:47–55.

8. Centers for Disease Control and Prevention National Center for Health Statistics. National Health and Nutrition Examination Survey. NHANES. https://wwwn.cdc.gov/nchs/nhanes/continuousnhanes/default.aspx?BeginYear=1999

9. Inker L, Eneanya ND, Coresh J, Tighiouart H, Wang D, Sang Y, Crews DC, Doria A, Estrella MM, Froissart M, Grams ME, Greene T, Grubb A, Gudnason V, Gutierrez OM, Kalil R, Karger AB, Mauer M, Navis G, Nelson RG, Poggio ED, Rodby R, Rossing P, Rule AD, Selvin E, Seegmiller JC, Shlipak MG, Torres VE, Yang W, Ballew SH, Couture SJ, Powe NR, Levey AS. New Creatinine- and Cystatin C–Based Equations to Estimate GFR without Race. New England Journal of Medicine 2021;385:1737–1749.

10. CKD Prognosis Consortium. Association of estimated glomerular filtration rate and albuminuria with all-cause and cardiovascular mortality in general population cohorts: a collaborative meta-analysis. Lancet 2010;375:2073–2081.

11. Kang M, Kwon S, Lee J, Shin J, Kim YC, Park JY, Bae E, Kim EY, Kim DK, Lim CS, Lee JP. Albuminuria within the Normal Range Can Predict All-Cause Mortality and Cardiovascular Mortality. Kidney360 2022;3:74–82.

12. Xu J, Knowler WC, Devereux RB, Yeh J, Umans JG, Begum M, Fabsitz RR, Lee ET. Albuminuria Within the ‘Normal’ Range and Risk of Cardiovascular Disease and Death in American Indians: The Strong Heart Study. American Journal of Kidney Diseases 2007;49:208–216.

13. Romundstad S, Holmen J, Kvenild K, Hallan H, Ellekjær H. Microalbuminuria and all-cause mortality in 2,089 apparently healthy individuals: A 4.4-year follow-up study. The Nord-Trøndelag Health Study (HUNT), Norway. American Journal of Kidney Diseases 2003;42:466–473.

14. Sung KC, Ryu S, Lee JY, Lee SH, Cheong E, Hyun YY, Lee KB, Kim H, Byrne CD. Urine Albumin/Creatinine Ratio Below 30 mg/g is a Predictor of Incident Hypertension and Cardiovascular Mortality. J Am Heart Assoc 2016;5:1–11.

15. Sung KC, Ryu S, Lee JY, Lee SH, Cheong E, Hyun YY, Lee KB, Kim H, Byrne CD. Urine Albumin/Creatinine Ratio Below 30 mg/g is a Predictor of Incident Hypertension and Cardiovascular Mortality. J Am Heart Assoc 2016;5:1–11.

16. Xu J, Knowler WC, Devereux RB, Yeh J, Umans JG, Begum M, Fabsitz RR, Lee ET. Albuminuria Within the ‘Normal’ Range and Risk of Cardiovascular Disease and Death in American Indians: The Strong Heart Study. American Journal of Kidney Diseases 2007;49:208–216.

17. Romundstad S, Holmen J, Kvenild K, Hallan H, Ellekjær H. Microalbuminuria and all-cause mortality in 2,089 apparently healthy individuals: A 4.4-year follow-up study. The Nord-Trøndelag Health Study (HUNT), Norway. American Journal of Kidney Diseases 2003;42:466–473.

18. Ärnlöv J, Evans JC, Meigs JB, Wang TJ, Fox CS, Levy D, Benjamin EJ, D’Agostino RB, Vasan RS. Low-grade albuminuria and incidence of cardiovascular disease events in nonhypertensive and nondiabetic individuals: The Framingham heart study. Circulation 2005;112:969–975.

19. Kidney Disease: Improving Global Outcomes (KDIGO) Blood Pressure Work Group. KDIGO 2021 Clinical Practice Guideline for the Management of Blood Pressure in Chronic Kidney Disease. Kidney Int 2021;99:S1–S87.

20. Mogensen CE. Microalbuminuria predicts clinical proteinuria and eraly mortality in maturity-onset diabetes. N Engl J Med 1984;310:356–360.

21. Greve S V., Blicher MK, Blyme A, Sehestedt T, Hansen TW, Rassmusen S, Vishram JKK, Ibsen H, Torp-Pedersen C, Olsen MH. Association between albuminuria, atherosclerotic plaques, elevated pulse wave velocity, age, risk category and prognosis in apparently healthy individuals. J Hypertens 2014;32:1034–1041.

22. Sehestedt T, Jeppesen J, Hansen TW, Rasmussen S, Wachtell K, Ibsen H, Torp-Pedersen C, Olsen MH. Thresholds for pulse wave velocity, urine albumin creatinine ratio and left ventricular mass index using SCORE, Framingham and ESH/ESC risk charts. J Hypertens 2012;30:1928–1936.

23. Heo NJ, Ahn JM, Lee TW, Chin HJ, Na KY, Chae DW, Kim S. Very low-grade albuminuria reflects susceptibility to chronic kidney disease in combination with cardiovascular risk factors. Hypertension Research 2010;33:573–578.

24. National Institute of Diabetes and Digestive and Kidney Diseases. 2020 USRDS Annual Data Report: Epidemiology of Kidney Disease in the United States. United States Renal Data System.

25. Ruilope LM, Ortiz A, Lucia A, Miranda B, Alvarez-llamas G, Barderas MG, Volpe M, Ruiz-hurtado G, Pitt B. Prevention of cardiorenal damage : importance of albuminuria. Eur Heart J 2022;00:1–12.

26. Deckert T, Feldt-Rasmussen B, Borch-Johnsen K, Kofoed-Enevoldsen A. Albuminuria reflects widespread vascular damage. The Steno hypothesis. Diabetologia 1989;32:219–226.

27. Salmon AHJ, Ferguson JK, Burford JL, Gevorgyan H, Nakano D, Harper SJ, Bates DO, Peti-Peterd J. Loss of the endothelial glycocalyx links albuminuria and vascular dysfunction. Journal of the American Society of Nephrology 2012;23:1339–1350.

28. Katz DH, Selvaraj S, Aguilar FG, Martinez EE, Beussink L, Kim KYA, Peng J, Sha J, Irvin MR, Eckfeldt JH, Turner ST, Freedman BI, Arnett DK, Shah SJ. Association of low-grade albuminuria with adverse cardiac mechanics: Findings from the hypertension genetic epidemiology network (hypergen) study. Circulation 2014;129:42–50.

29. Boorsma EM, Maaten JM ter, Damman K, Essen BJ van, Zannad F, Veldhuisen DJ van, Samani NJ, Dickstein K, Metra M, Filippatos G, Lang CC, Ng L, Anker SD, Cleland JG, Pellicori P, Gansevoort RT, Heerspink HJL, Voors AA, Emmens JE. Albuminuria as a marker of systemic congestion in patients with heart failure. Eur Heart J 2022;44:368–380.

30. Martens RJH, Houben AJHM, Kooman JP, Berendschot TTJM, Dagnelie PC, Kallen CJH Van Der, Kroon AA, Leunissen KML, Sande FM Van Der, Schaper NC, Schouten JSAG, Schram MT, Sep SJS, Sörensen BM, Henry RMA, Stehouwer CDA. Microvascular endothelial dysfunction is associated with albuminuria: The Maastricht Study. J Hypertens 2018;36:1178–1187.

31. Klausen K, Borch-Johnsen K, Feldt-Rasmussen B, Jensen G, Clausen P, Scharling H, Appleyard M, Jensen JS. Very low levels of microalbuminuria are associated with increased risk of coronary heart disease and death independently of renal function, hypertension, and diabetes. Circulation 2004;110:32–35.

32. Tanaka F, Komi R, Makita S, Onoda T, Tanno K, Ohsawa M, Itai K, Sakata K, Omama S, Yoshida Y, Ogasawara K, Ishibashi Y, Kuribayashi T, Okayama A, Nakamura M. Low-grade albuminuria and incidence of cardiovascular disease and all-cause mortality in nondiabetic and normotensive individuals. J Hypertens 2016;34:506–512.

33. Allin KH, Bojesen SE, Nordestgaard BG. Baseline C-reactive protein is associated with incident cancer and survival in patients with cancer. Journal of Clinical Oncology 2009;27:2217–2224.

34. Jørgensen L, Heuch I, Jenssen T, Jacobsen BK. Association of albuminuria and cancer incidence. Journal of the American Society of Nephrology 2008;19:992–998.

35. Banerjee T, Crews D, Wesson DE, Dharmarajan S, Saran R, Ríos Burrows N, Saydah S, Powe NR, Hsu C yuan, Bibbins-Domingo K, McCulloch C, Grubbs V, Peralta C, Shlipak M, Rubinsky A, Hsu R, Coresh J, Tuot D, Steffick D, Gillespie B, Herman W, Port F, Robinson B, Shahinian V, Yee J, Young E, McClellan W, O’Hare A, Fava M, Tilea A, Williams D, Eberhardt M, Flowers N, Geiss L, Jordan R, Mondesire J, Moore B, Myers G, Pavkov M, Rolka D, Schoolwerth A, Valdez R, Waller L. Food Insecurity, CKD, and Subsequent ESRD in US Adults. American Journal of Kidney Diseases 2017;70:38–47.

36. Crews DC, Kuczmarski MF, Grubbs V, Hedgeman E, Shahinian VB, Evans MK, Zonderman AB, Burrows NR, Williams DE, Saran R, Powe NR, Centers for Disease Control and Prevention Chronic Kidney Disease Surveillance Team for the C for DC and PCKDS. Effect of food insecurity on chronic kidney disease in lower-income Americans. Am J Nephrol 2014;39:27–35.

37. Crews DC, Novick TK. Social Determinants of CKD Hotspots. Seminars in Nephrology.

38. Golovaty I, Tien PC, Price JC, Sheira L, Seligman H, Weiser SD. Food Insecurity May Be an Independent Risk Factor Associated with Nonalcoholic Fatty Liver Disease among Low-Income Adults in the United States. Journal of Nutrition 2020;150:91–98.

39. Asselbergs FW, Harst P Van Der, Gilst WH Van. Letter regarding article by Arnlov et al, ‘Low-grade albuminuria and incidence of cardiovascular disease events in nonhypertensive and nondiabetic individuals’ [1]. Circulation 2006;113:1–2.

40. Selvin E, Juraschek SP, Eckfeldt JH, Levey AS, Inker LA, Coresh J. Within-Person Variability in Kidney Measures. Am J Kidney Dis 2013;61:716–722.

41. Waikar S, Rebholz CM, Zheng Z, Hurwitz S, Hsu C-Y, Feldman HI, Xie D, Liu KD, Mifflin TE, Eckfeldt JH, Kimmel PL, Vasan RS, Bonventre J V., Inker LA, Coresh J, Chronic Kidney Disease Biomarkers

42. Consortium Investigators. Biological Variability of Estimated GFR and Albuminuria in CKD. Am J Kidney Dis 2018;72:538–546.

42. Zanoli L, Empana JP, Perier MC, Alivon M, Ketthab H, Castellino P, Laude D, Thomas F, Pannier B, Laurent S, Jouven X, Boutouyrie P. Increased carotid stiffness and remodelling at early stages of chronic kidney disease. J Hypertens 2019;37:1176–1182.

43. Visseren FLJ, MacH F, Smulders YM, Carballo D, Koskinas KC, Bäck M, Benetos A, Biffi A, Boavida JM, Capodanno D, Cosyns B, Crawford C, Davos CH, Desormais I, Angelantonio E DI, Franco OH, Halvorsen S, Hobbs FDR, Hollander M, Jankowska EA, Michal M, Sacco S, Sattar N, Tokgozoglu L, Tonstad S, Tsioufis KP, DIs I Van, Gelder IC Van, Wanner C, Williams B, Backer G De, Regitz-Zagrosek V, Aamodt AH, Abdelhamid M, Aboyans V, Albus C, Asteggiano R, Bäck M, Borger MA, Brotons C, Ielutkienė J, Cifkova R, Cikes M, Cosentino F, Dagres N, Backer T De, Bacquer Di De, Delgado V, Ruijter H Den, Dendale P, Drexel H, Falk V, Fauchier L, Ference BA, Ferrières J, Ferrini M, Fisher M, Fliser D, Fras Z, Gaita D, Giampaoli S, Gielen S, Graham I, Jennings C, Jorgensen T, Kautzky-Willer A, Kavousi M, Koenig W, Konradi A, Kotecha Di, Landmesser U, Lettino M, Lewis BS, Linhart A, Løchen ML, Makrilakis K, Mancia G, Marques-Vidal P, McEvoy JW, McGreavy P, Merkely B, Neubeck L, Nielsen JC, Perk J, Petersen SE, Petronio AS, Piepoli M, Pogosova NG, Prescott EIB, Ray KK, Reiner Z, Richter DiJ, Rydén L, Shlyakhto E, Sitges M, Sousa-Uva M, Sudano I, Tiberi M, Touyz RM, Ungar A, Verschuren WMM, Wiklund O, Wood D, Zamorano JL, Smulders YM, Carballo D, Koskinas KC, Bäck M, Benetos A, Biffi A, Boavida JM, Capodanno D, Cosyns B, Crawford CA, Desormais I, Franco Duran OH, Halvorsen S, Richard Hobbs FD, Hollander M, Jankowska EA, Michal M, Sattar N, Tokgozoglu L, Tonstad S, DIs I Van, Gelder IC Van, Williams B. 2021 ESC Guidelines on cardiovascular disease prevention in clinical practice. Eur Heart J 2021;42:3227– 3337.

