## Supplemental Tables for "The relationship between low levels of albuminuria and cardiovascular mortality among apparently healthy adults"

| Table S1. Missingness of key variables (N=16,247) | |
|---|---|
| | N(%) |
| Age | 0 |
| Female sex | 0 |
| Race or Ethnicity | 0 |
| Education | 20 (0.1) |
| Income | 1234 (6.2) |
| Health insurance | 71 (0.4) |
| Food insecurity | 327 (2.0) |
| Smoking status | 14 (0.1) |
| Systolic blood pressure | 890 (5.5) |
| Diastolic blood pressure | 904 (5.6) |
| Hemoglobin A1c | 28 (0.2) |
| Body mass index | 146 (0.9) |
| Total cholesterol | 9 (0.1) |
| eGFR | 0 |
| Statin use | 0 |

**Table S2. Clinical correlates of albuminuria.**

| | Univariate | | | Multivariable | | |
|---|---|---|---|---|---|---|
| | **β Coefficient (95% CI)** | **% Change (95% CI)** | ***p*-value** | **β Coefficient (95% CI)** | **% Change (95% CI)** | ***p*-value** |
| 10-year increase in age | 0.03 (0.03, 0.04) | 3.5 (2.6, 4.4) | <0.0001 | 0.09 (0.08, 0.1) | 9.9 (8.6, 11.2) | <0.0001 |
| Male sex | -0.3 (-0.3, -0.3) | -26.7 (-28.3, -25.1) | <0.0001 | -0.4 (-0.4, -0.4) | -31.6 (-33.2, -29.9) | <0.0001 |
| Race or Ethnicity | | | | | | |
| Non-Hispanic White | [Ref] | [Ref] | | [Ref] | [Ref] | |
| Mexican | 0.06 (0.02, 0.09) | 5.8 (2.1, 9.5) | 0.002 | -0.01 (-0.05, 0.03) | -1.2 (-4.9, 2.6) | 0.52 |
| Other Hispanic | 0.009 (-0.03, 0.05) | 0.9 (-3.2, 5.3) | 0.66 | -0.04 (-0.09, 0.0005) | -4.2 (-8.2, 0.05) | 0.05 |
| Non-Hispanic Black | -0.07 (-0.1, -0.03) | -6.5 (-9.8, -3.2) | 0.002 | -0.06 (-0.09, -0.02) | -5.7 (-9.0, -2.3) | 0.001 |
| Other | 0.03 (-0.02, 0.07) | 2.6 (-1.9, 7.2) | 0.26 | -0.001 (-0.05, 0.04) | -0.1 (-4.5, 4.5) | 0.97 |
| Education | | | | | | |
| Below High School | 0.05 (0.02, 0.09) | 5.6 (2.4, 8.9) | 0.0007 | 0.01 (-0.02, 0.05) | 1.2 (-2.3, 4.8) | 0.50 |
| High School / GED | 0.008 (-0.02, 0.04) | 0.8 (-2.1, 3.8) | 0.60 | -0.004 (-0.03, 0.02) | -0.4 (-3.2, 2.4) | 0.76 |
| Above High School | [Ref] | [Ref] | | [Ref] | [Ref] | |
| Uninsured | 0.02 (-0.01, 0.04) | 1.6 (-1.1, 4.4) | 0.25 | 0.04 (0.008, 0.07) | 3.7 (0.8, 6.7) | 0.01 |
| Food insecurity | 0.06 (0.03, 0.08) | 5.7 (2.7, 8.8) | 0.0002 | 0.05 (0.02, 0.08) | 4.9 (1.6, 8.3) | 0.003 |
| Current/former smoking | -0.01 (-0.03, 0.01) | -1.0 (-3.1, 1.1) | 0.33 | -- | -- | |
| 5kg/m$^2$ increase in BMI | -0.02 (-0.03, -0.007) | -1.7 (-2.7, -0.7) | 0.001 | -0.02 (-0.04, -0.01) | -2.4 (-3.5, -1.4) | <0.0001 |
| 1% increase in hemoglobin A1c | 0.08 (0.06, 0.1) | 8.9 (5.7, 12.1) | <0.0001 | 0.09 (0.06, 0.1) | 9.5 (5.8, 13.2) | <0.0001 |
| 1mg/dL increase in serum albumin | -0.004 (-0.008, -0.0005) | -0.4 (-0.8, -0.05) | 0.03 | 0.02 (0.02, 0.02) | 2.0 (1.5, 2.4) | <0.0001 |
| 50mg/dL increase in total cholesterol | -0.004 (-0.02, 0.01) | -0.4 (-1.9, 1.0) | 0.55 | -0.03 (-0.04, -0.01) | -2.8 (-4.2, -1.4) | 0.0002 |
| 10ml/min decrease in eGFR | -0.03 (-0.04, -0.03) | -3.4 (-4.1, -2.6) | <0.0001 | -0.08 (-0.09, -0.07) | -7.4 (-8.3, -6.5) | <0.0001 |
| 10mmHg increase in SBP | -0.02 (-0.03, -0.002) | -1.7 (-3.2, -0.2) | 0.03 | 0.03 (0.02, 0.05) | 3.1 (1.5, 4.7) | 0.0002 |
| 5mmHg increase in DBP | -0.007 (-0.01, 0.001) | -0.7 (-1.5, 0.1) | 0.09 | -- | -- | |

The dependent variable in these models is log-transformed urine albumin concentration. All models are adjusted for log-transformed urine creatinine concentration. Smoking status and DBP not included in multivariable regression as $p$ >0.10 when added to the model. Percent change reflects the exponentiated β coefficient.

Abbreviations: BMI, body mass index; CI, confidence interval; DBP, diastolic blood pressure; eGFR, estimated glomerular filtration rate; GED, general education diploma; SBP, systolic blood pressure.

| Table S3. Association between albuminuria and non-cardiovascular mortality. | | | | | |
|---|---|---|---|---|---|
| | **UACR, mg/g** | | **Tertile 1 UACR < 4.3 mg/g** | **Tertile 2 UACR 4.3-7.1 mg/g** | **Tertile 3 UACR > 7.1 mg/g** |
| | **HR (95% CI)** | ***p*-value** | **HR (95% CI)** | **HR (95% CI)** | **HR (95% CI)** |
| **Non-Cardiovascular mortality** | | | | | |
| No. of events | 777 | | 189 | 249 | 339 |
| Unadjusted | 1.35 (1.21, 1.50) | <0.0001 | Reference | 1.32 (1.03, 1.70) | 1.75 (1.37, 2.24) |
| Model 1 | 1.24 (1.12, 1.38) | 0.0001 | Reference | 1.25 (0.96, 1.62) | 1.46 (1.13, 1.89) |
| Model 2 | 1.28 (1.15, 1.43) | <0.0001 | Reference | 1.35 (1.03, 1.76) | 1.57 (1.21, 2.03) |
| Model 3 | 1.26 (1.13, 1.41) | <0.0001 | Reference | 1.31 (1.00, 1.70) | 1.51 (1.17, 1.95) |

Model 1: adjusted for age, sex, race or ethnicity, insurance, smoking, survey year, BMI
Model 2: Model 1 + hemoglobin A1c, total cholesterol, systolic blood pressure, serum albumin, statin use
Model 3: Model 2 + eGFR

| **Table S4. The association between albuminuria and mortality.** | | | | |
|---|---|---|---|---|
| | **UACR ≥ 7 mg/g (vs < 7 mg/g)** | | **UACR per 10 mg/g increase** | |
| | **HR (95% CI)** | ***p*-value** | **HR (95% CI)** | ***p*-value** |
| **Cardiovascular mortality** | | | | |
| No. of events | 177 | | 177 | |
| Unadjusted | 2.79 (2.00, 3.91) | <0.0001 | 2.02 (1.65, 2.48) | <0.0001 |
| Model 1 | 2.01 (1.42. 2.86) | 0.0001 | 1.57 (1.25, 1.97) | <0.0001 |
| Model 2 | 2.19 (1.56, 3.09) | <0.0001 | 1.68 (1.34, 2.10) | <0.0001 |
| Model 3 | 2.13 (1.50, 3.01) | <0.0001 | 1.64 (1.31, 2.06) | <0.0001 |
| **Non-Cardiovascular mortality** | | | | |
| No. of events | 777 | | 777 | |
| Unadjusted | 1.49 (1.24, 1.79) | <0.0001 | 1.67 (1.42, 1.91) | <0.0001 |
| Model 1 | 1.28 (1.05, 1.55) | 0.01 | 1.47 (1.28, 1.70) | <0.0001 |
| Model 2 | 1.32 (1.08, 1.62) | 0.008 | 1.55 (1.33, 1.80) | <0.0001 |
| Model 3 | 1.29 (1.05, 1.58) | 0.02 | 1.52 (1.31, 1.77) | <0.0001 |
| **All-cause mortality** | | | | |
| No. of events | 954 | | 954 | |
| Unadjusted | 1.66 (1.42, 1.95) | <0.0001 | 1.73 (1.53, 1.96) | <0.0001 |
| Model 1 | 1.39 (1.18, 1.64) | 0.0001 | 1.50 (1.33, 1.69) | <0.0001 |
| Model 2 | 1.45 (1.22, 1.72) | <0.0001 | 1.58 (1.39, 1.79) | <0.0001 |
| Model 3 | 1.41 (1.19, 1.68) | 0.0001 | 1.55 (1.36, 1.76) | <0.0001 |
| Model 1: adjusted for age, sex, race or ethnicity, insurance, smoking, survey year, BMI<br>Model 2: Model 1 + hemoglobin A1c, total cholesterol, systolic blood pressure, serum albumin, statin use<br>Model 3: Model 2 + eGFR | | | | |

| **Table S5. Tests for effect modification** | |
|---|---|
| | ***p*-value** |
| **Cardiovascular mortality** | |
| Male sex | 0.29 |
| Race or Ethnicity | |
| Non-Hispanic White | [Referent] |
| Mexican American | 0.78 |
| Other Hispanic | 0.23 |
| Non-Hispanic Black | 0.69 |
| Other Race | 0.60 |
| Hemoglobin A1c | 0.77 |
| BMI | 0.26 |
| **Non-Cardiovascular Mortality** | |
| Male sex | 0.79 |
| Race or Ethnicity | |
| Non-Hispanic White | [Referent] |
| Mexican American | 0.72 |
| Other Hispanic | 0.60 |
| Non-Hispanic Black | 0.37 |
| Other Race | 0.68 |
| Hemoglobin A1c | 0.98 |
| BMI | 0.03 |
| **All-cause mortality** | |
| Male sex | 0.82 |
| Race or Ethnicity | |
| Non-Hispanic White | [Referent] |
| Mexican American | 0.89 |
| Other Hispanic | 0.65 |
| Non-Hispanic Black | 0.50 |
| Other Race | 0.53 |
| Hemoglobin A1c | 0.81 |
| BMI | 0.01 |
| Adjusted for: age, sex, race or ethnicity, insurance, smoking, survey year, BMI, hemoglobin A1c, total cholesterol, systolic blood pressure, serum albumin, statin use, eGFR | |
